# Association between food environments and fetal growth in pregnant Brazilian women

**DOI:** 10.1101/2022.08.24.22279156

**Authors:** Audêncio Victor, Ana Raquel Manuel Gotine, Ila R. Falcão, Andrea Ferreira, Renzo Flores-Ortiz, Sancho Pedro Xavier, Melsequisete Daniel Vasco, Natanael de Jesus Silva, Manuel Mahoche, Osiyallê Akanni Silva Rodrigues, Rita da Cassia Ribeiro, Patrícia H. Rondó, Maurício L. Barreto

## Abstract

**Introduction:** Birth weight is described as one of the main determinants of newborns’ chances of survival. Among the associated causes, or risk factors, the mother’s nutritional status strongly influences fetal growth and birth weight outcomes of the concept. This study evaluates the association between food deserts, small for gestational age (SGA), large for gestational age (LGA) and low birth weight (LBW) newborns.

**Design:** This is a cross-sectional population study, resulting from individual data from the Live Birth Information System (SINASC), and municipal data from mapping food deserts (CAISAN) in Brazil. The newborn’s size was defined as follows: appropriate for gestational age (between 10^th^ and 90^th^ percentile), SGA (<10^th^ percentile), LGA (>90^th^ percentile), and low birth weight <2,500g. To characterize food environments, we used tertiles of the density of establishments which sell *in natura* and ultra-processed foods. Logistic regression modeling was conducted to investigate the associations of interest.

**Results:** We analyzed 2,632,314 live births in Brazil in 2016. Following due adjustment, it was observed that women from municipalities in which there was a limited supply of *in natura* foods presented a higher chance of a SGA [OR_2nd tertile_:1.06 (1.05-1.07)] and LBW [OR_2nd tertile_: 1.11(1.09-1.12)] newborn. On the other hand, municipalities in which there was a greater supply of ultra-processed foods presented higher chances for a SGA [OR_3rd tertile_:1.04 (1.02-1.06)] and LBW [OR_2nd tertile_:1.13 (1.11-1.16)] newborn. Stratification showed that Black and Mixed/Brown women were associated with SGA [OR_3rd tertile_: 1.09 (1.01-1.18)] and [OR_3rdtertile_:1.06 (1.04-1.09)], respectively, and mixed-race women were also associated with LBW [OR_3rd tertile_:1.17 (1.14-1.20)], while indigenous women were associated with LGA [OR_3rd tertile_: 1.20(1.01-1.45)].

**Conclusions:** Living in areas with limited access to healthy foods was associated with an increased chance of SGA and low weight newborns, especially between Black and Mixed/Brown women. Initiatives focused on minimizing the effects of these food environments, and which aim to reduce social inequalities, are urgently required in the Brazilian context.

## Introduction

Food deserts is a recent term used to indicate the lack of availability of healthy foods (whether in quantity or quality) in urban or rural areas, usually of a low income^1^. Low economic conditions limit access to healthy foods, thereby contributing towards the acquisition of processed and ultra-processed foods, which contribute to a gain in excessive gestational weight, which has been associated with the birth of large for gestational age (LGA) babies^2^.

Among the various ways that food environments affect health and quality of life, we highlight the impact of food deserts, and the consequent food insecurity in deprived communities, inserted in unhealthy global macroeconomic and industrial policies^2^. In addition to impacts on the general population, food deserts also profoundly affect babies’ lives, from their intrauterine growth to conception, and their consequent growth and development^1^.

Healthy food consumption is essential during the pre-conceptional period, pregnancy, and breastfeeding, to guarantee maternal-fetal health and the concept. In the configuration of a maternal diet, it is important to consider that the inadequate intake of vitamins and minerals, and high consumption of food and drinks with refined sugar and saturated fat, increase the occurrence of low birth weight (LBW) babies^4^. During the 20^th^ century, high risk newborns were defined as those with a weight of under 2,500g, also known as low birth weight (LBW).

Maternal body composition during intrauterine growth may contribute towards a greater risk of cardiometabolic disease in small or large for gestational age (SGA and LGA) newborns. The compensations of intrauterine growth through recovery, or reduction, of postnatal growth may lead to adverse consequences^7^. Being born SGA involves an increase in infant morbimortality rates, and is also related to alterations in the growth pattern and body composition, which may be associated with the development of risk factors connected to metabolic and cardiovascular diseases^8^. The gain of excessive gestational weight and the presence of metabolic comorbidities is one of the maternal factors associated with the birth of LGA babies, while insufficient gestational weight gain is associated with the birth of SGA babies^9,10^. Birth weight is described as the main determinant of the chances of newborns’ survival, and has been significantly associated with various factors, such as the mother’s level of education, the number of prenatal appointments, ethnicity/race, and maternal weight during pregnancy^11,12^, including food deserts^13^.

It is vital to understand food deserts and their various outlines in important outcomes of interest to public health in this context^14^, such as the association between food deserts and the birth of SGA, LGA, and LBW babies.

## Methods

### Study design, population, and data source

This is a cross-sectional population study resulting from merging data from the Ministry of Health Unified Health System, Department of Information and IT (DATASUS) 2016 Live Birth Information System (SINASC), obtained by the Data Science Applied to Health Platform (PCDaS/Icict/Fiocruz)^15^, and food desert data produced by the <u>Interministerial Chamber of Food and Nutritional</u> Security (CAISAM), at the municipal level (aggregate) in 2016^16^.

### Study variables

#### Outcome: Fetal growth

Fetal growth was categorized as: appropriate for gestational age (between 10th and 90^th^ percentile), SGA (<10^th^ percentile), or LGA (>90^th^ percentile), using specific curves by sex corresponding to single live births, as established by the INTERGROWTH-21^st^ Consortium^17^, to classify weight by gestational age (24/0 to 42/0 gestational weeks).

The LBW outcome was considered from the dichotomous classification of the birth weight variable: LBW: < 2,500 g, and normal weight: ≥ 2,500 g.

### Main independent variables

The independent variables were subdivided into two levels: individual and context. The main independent variable was the food environment context at the municipal level.

Food environment information was obtained through a study conducted by the Interministerial chamber of Food and Nutritional Security (CAISAN) (CAISAN, 2018). This study proposed mapping food deserts in Brazil. Thus, this purpose used information from the Annual Social Information Report (RAIS-2016) (only the establishment database). In this database, the establishments are classified following the National Classification of Economic Activities (CNAE – an instrument for the national standardization of economic activity codes, and the qualifying criteria used by various tax administration bodies in the country). Data from the organic fair map of street markets produced by the Brazilian Institute for Consumer Defense (IDEC), SAN map of markets, and food markets listed on the electronic sites of city halls in Brazilian state capitals were also incorporated mapping. With the establishments located, we then moved on to analyze what the population acquires at each category of establishments. Thus, the database selected was Family Budget Research (POF 2008-2009). Food acquired by the population, and the respective locations for these purchases can be identified on this database. The food acquired was then classified following the four categories proposed by the 2014 Food Guide for the Brazilian Population (Ministry of Health, 2014). The next step was connecting the purchase locations reported in the POF with the establishments classified by the CNAE. Having completed the previous stages, we determined the acquisition percentage of each food category in the guide by the CNAE. Analysis of the establishment profiles and RAIS establishment classification were the next steps. Thus, the establishments were classified as: a) establishments that predominantly (more than 50%) sell *in natura* food; b) ultra-processed food, and c) mixed establishments, where there was no predominance of supplying healthy or unhealthy foods.

To characterize the food environments, we only used the density of the establishments which sold *in natura* and ultra-processed foods (per 10,000 inhabitants). These results were used in the statistical models in tertiles.

### Covariates

The covariates considered at the individual level were as follows: mother’s age (≤17; 18-19; 20-34; and ≥35); marital status (married, civil partnership, single, widow, or separated); the mother’s level of education at time of birth (none; 1-7; 8-11; and 12 or more); gestational age in weeks (22-27, 28-31, 32-36, and 37-42 weeks), number of prenatal appointments (none, between 1 and 3, between 4 and 6, and 7 and more); newborn’s sex (male or female); and mother’s race/color (White; Black, Yellow (Asian descendant), Mixed/Brown, and indigenous).

The contextual variables include: (Gross Domestic Product (GDP) per capita, Gini index, unemployment rate, Family Health Strategy (ESF) coverage, municipal Human Development Index obtained from IBGE 2000 and 2010 Demographic Census data, extrapolated for 2016, and provided by the IBGE SIDRA Automatic Recovery^18^ system by the DATASUS TabNet tool^19^, and the United Nations Development Programme (UNDP) Human Development Atlas in Brazil, ^20^.

### Statistical analysis

The socio-economic, maternal, and live birth characteristics were summarized through frequency distributions. Investigation of the food deserts associated with the baby’s size, was conducted using logistic regression models, to investigate factors associated with SGA, LGA, LBW, and food deserts, defined by the density of *in natura* and ultra-processed foods (in tertiles), which formed the main independent variables. The models were simultaneously adjusted by the covariates. The variables were included step-by-step, and we tested those which presented p <0.25 in the model, in the univariate analysis. The magnitude was quantified through the odds ratio (OR), with a 95% confidence interval (CI 95%).

All the analyses were conducted using the available covariables, which were considered plausible and relevant in literature^21–23^. In order to select the contextual variables and the theoretical framework, correlation analysis was conducted to evaluate multicollinearity. A conceptual model was adopted to introduce the variables **(Figure 2)**.

The analysis was carried out using R, version 3.6.1 (http://www.r-project.org), and RStudio software, version 1.2.1335.

### Ethical issues

Secondary, individual, anonymized, and aggregated public domain data at the municipal level was used exclusively. Therefore, free and informed consent and approval by the Ethics and Research Committee were exempted, following the National Health Council in Brazil, National Commission for Research Ethics Resolution Nº 466/2012.

## Results

In relation to the sociodemographic data, Brazil’s GDP per capita of BRL 17,600 was observed. The unemployment rate was approximately 4.6%. The percentage of Family Health Strategy (ESF) coverage, the main basic health care policy in Brazil, was 88.14%. Further information can be found in **Table 1**.

2,632,314 live births were included in the study, 186,206 (7.07 %), 428,972 (16.30 %), and 188,450 (7.16 %) of which were classified as SGA, LGA, and LBW, respectively **(Figure 1)**.

**Figure 1.**
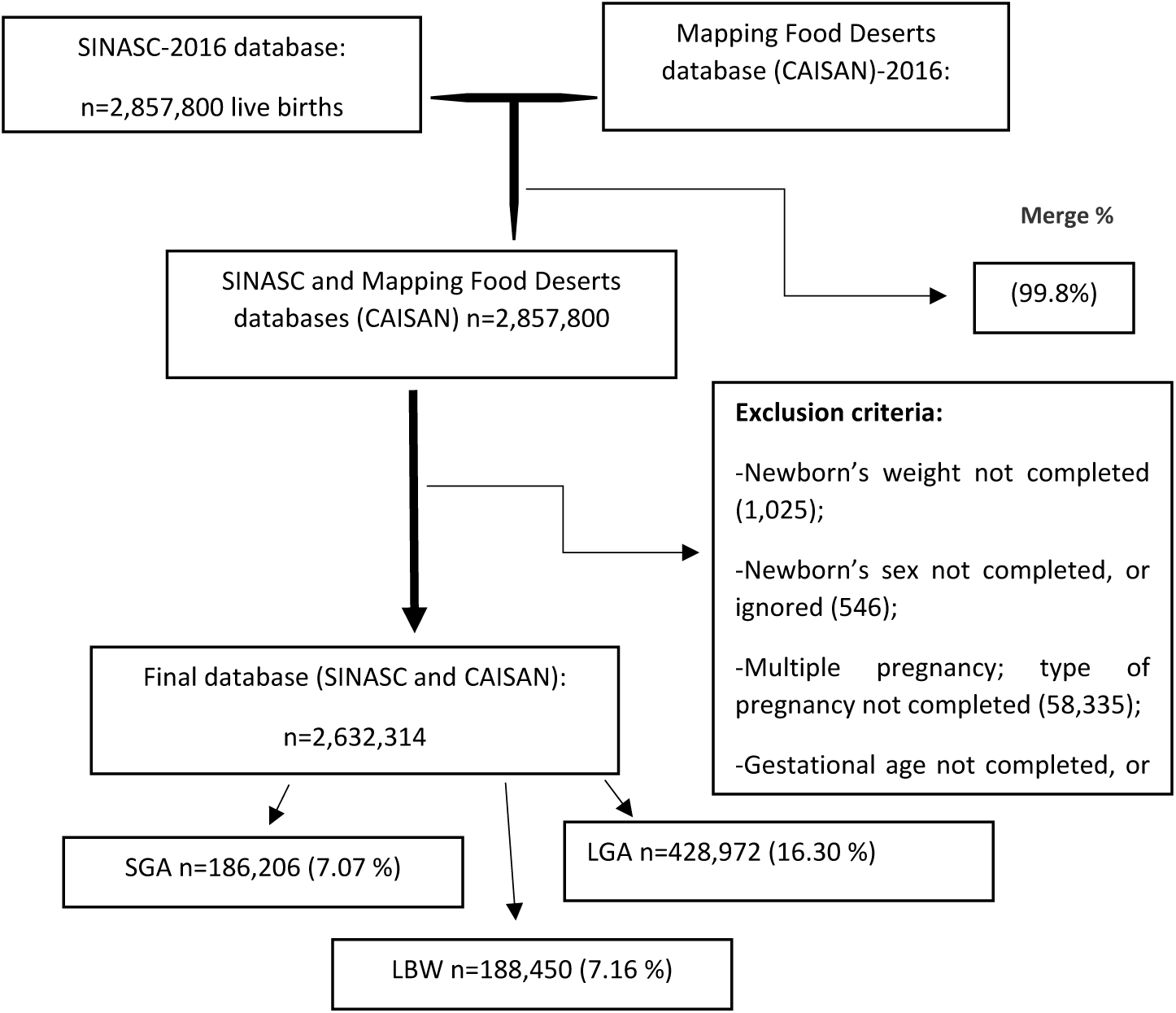
Study population.

With regards to the study population characteristics, it was noted that SGA and LBW are more common in women aged ≥35, with fewer years of education, indigenous and black mothers, with a lower number of prenatal appointments, single, widows, or divorced, and with female newborns (Table 2).

**Table 2.**
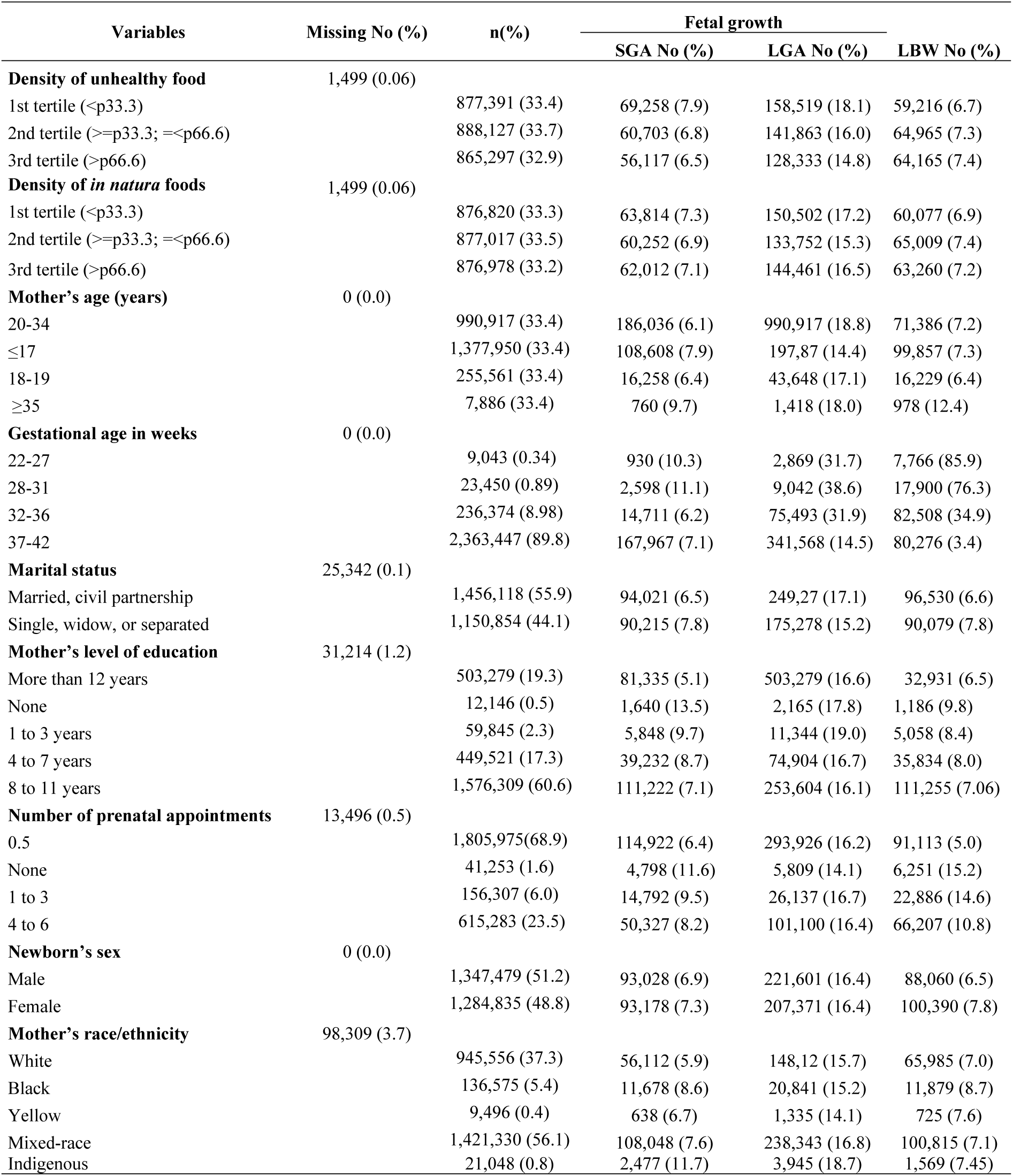
Food environment, socio-economic, maternal, and live birth characteristics, in accordance with fetal growth and birth weight (n=2,632,314) in 2016.

| Variables | Missing No (%) | n(%) | Fetal growth |  |  |
| --- | --- | --- | --- | --- | --- |
|  |  |  | SGA No (%) | LGA No (%) | LBW No (%) |
| **Density of unhealthy food** | 1,499 (0.06) |  |  |  |  |
| 1st tertile (<p33.3) |  | 877,391 (33.4) | 69,258 (7.9) | 158,519 (18.1) | 59,216 (6.7) |
| 2nd tertile (>=p33.3; <=p66.6) |  | 888,127 (33.7) | 60,703 (6.8) | 141,863 (16.0) | 64,965 (7.3) |
| 3rd tertile (>p66.6) |  | 865,297 (32.9) | 56,117 (6.5) | 128,333 (14.8) | 64,165 (7.4) |
| **Density of *in natura* foods** | 1,499 (0.06) |  |  |  |  |
| 1st tertile (<p33.3) |  | 876,820 (33.3) | 63,814 (7.3) | 150,502 (17.2) | 60,077 (6.9) |
| 2nd tertile (>=p33.3; <=p66.6) |  | 877,017 (33.5) | 60,252 (6.9) | 133,752 (15.3) | 65,009 (7.4) |
| 3rd tertile (>p66.6) |  | 876,978 (33.2) | 62,012 (7.1) | 144,461 (16.5) | 63,260 (7.2) |
| **Mother's age (years)** | 0 (0.0) |  |  |  |  |
| 20-34 |  | 990,917 (33.4) | 186,036 (6.1) | 990,917 (18.8) | 71,386 (7.2) |
| ≤17 |  | 1,377,950 (33.4) | 108,608 (7.9) | 197,87 (14.4) | 99,857 (7.3) |
| 18-19 |  | 255,561 (33.4) | 16,258 (6.4) | 43,648 (17.1) | 16,229 (6.4) |
| ≥35 |  | 7,886 (33.4) | 760 (9.7) | 1,418 (18.0) | 978 (12.4) |
| **Gestational age in weeks** | 0 (0.0) |  |  |  |  |
| 22-27 |  | 9,043 (0.34) | 930 (10.3) | 2,869 (31.7) | 7,766 (85.9) |
| 28-31 |  | 23,450 (0.89) | 2,598 (11.1) | 9,042 (38.6) | 17,900 (76.3) |
| 32-36 |  | 236,374 (8.98) | 14,711 (6.2) | 75,493 (31.9) | 82,508 (34.9) |
| 37-42 |  | 2,363,447 (89.8) | 167,967 (7.1) | 341,568 (14.5) | 80,276 (3.4) |
| **Marital status** | 25,342 (0.1) |  |  |  |  |
| Married, civil partnership |  | 1,456,118 (55.9) | 94,021 (6.5) | 249,27 (17.1) | 96,530 (6.6) |
| Single, widow, or separated |  | 1,150,854 (44.1) | 90,215 (7.8) | 175,278 (15.2) | 90,079 (7.8) |
| **Mother's level of education** | 31,214 (1.2) |  |  |  |  |
| More than 12 years |  | 503,279 (19.3) | 81,335 (5.1) | 503,279 (16.6) | 32,931 (6.5) |
| None |  | 12,146 (0.5) | 1,640 (13.5) | 2,165 (17.8) | 1,186 (9.8) |
| 1 to 3 years |  | 59,845 (2.3) | 5,848 (9.7) | 11,344 (19.0) | 5,058 (8.4) |
| 4 to 7 years |  | 449,521 (17.3) | 39,232 (8.7) | 74,904 (16.7) | 35,834 (8.0) |
| 8 to 11 years |  | 1,576,309 (60.6) | 111,222 (7.1) | 253,604 (16.1) | 111,255 (7.06) |
| **Number of prenatal appointments** | 13,496 (0.5) |  |  |  |  |
| 0.5 |  | 1,805,975(68.9) | 114,922 (6.4) | 293,926 (16.2) | 91,113 (5.0) |
| None |  | 41,253 (1.6) | 4,798 (11.6) | 5,809 (14.1) | 6,251 (15.2) |
| 1 to 3 |  | 156,307 (6.0) | 14,792 (9.5) | 26,137 (16.7) | 22,886 (14.6) |
| 4 to 6 |  | 615,283 (23.5) | 50,327 (8.2) | 101,100 (16.4) | 66,207 (10.8) |
| **Newborn's sex** | 0 (0.0) |  |  |  |  |
| Male |  | 1,347,479 (51.2) | 93,028 (6.9) | 221,601 (16.4) | 88,060 (6.5) |
| Female |  | 1,284,835 (48.8) | 93,178 (7.3) | 207,371 (16.4) | 100,390 (7.8) |
| **Mother's race/ethnicity** | 98,309 (3.7) |  |  |  |  |
| White |  | 945,556 (37.3) | 56,112 (5.9) | 148,12 (15.7) | 65,985 (7.0) |
| Black |  | 136,575 (5.4) | 11,678 (8.6) | 20,841 (15.2) | 11,879 (8.7) |
| Yellow |  | 9,496 (0.4) | 638 (6.7) | 1,335 (14.1) | 725 (7.6) |
| Mixed-race |  | 1,421,330 (56.1) | 108,048 (7.6) | 238,343 (16.8) | 100,815 (7.1) |
| Indigenous |  | 21,048 (0.8) | 2,477 (11.7) | 3,945 (18.7) | 1,569 (7.45) |

**Table 3** presents the results of the adjusted binary logistics regression, to evaluate the association between densities of processed and *in natura* foods, and birth weight. Following due adjustments, it was observed that women from municipalities where there was a limited supply of *in natura* foods presented a higher chance of a SGA newborn [OR_2nd tertile_:1.06 (1.05-1.07)]; LBW [OR_1st tertile_: 1.10 (1.08-1.12), OR_2nd tertile_: 1.11(1.09-1.12) dose-response effect]; stratification demonstrated that being of Mixed/Brown-race was associated with SGA [OR_1st tertile:_1.02 (1.01-1.03), OR_2nd tertile:_ 1.07 (1.06-1.10) doseresponse effect], LBW [OR_1st_ tertile:1.13 (1.12-1.16), OR_2nd tertile_: 1.12 (1.10-1.14)], and Black with LBW [OR_1st tertile:_ 1.14 (1.07-1.22)]. In parallel, municipalities where there was a higher supply of ultra-processed foods, presented greater chances of a SGA [OR_3rd tertile_:1.04 (1,02-1.06)] and LBW newborn [OR_1st tertile_: 1.11(1.09-1.13), OR_2nd tetcile_:1.13 (1.11-1.16) dose-response effect]. Stratification showed that black and mixed-race women, respectively: were associated with SGA [OR_3rd tertile_: 1.09 (1.01-1.18)] and [OR_3rd tertile_:1.06 (1.04-1.09)]. Only mixed-race women were associated with LBW [OR_2nd tertile_:1.09 (1.07-1.12), OR_3rd tertile_:1.17 (1.14-1.20)]. Being indigenous was associated with LGA [OR_3rd tertile_: 1.20(1.01-1.45)]. Raw analyses are provided in the supplementary tables (Table S1).

**Table 3.**
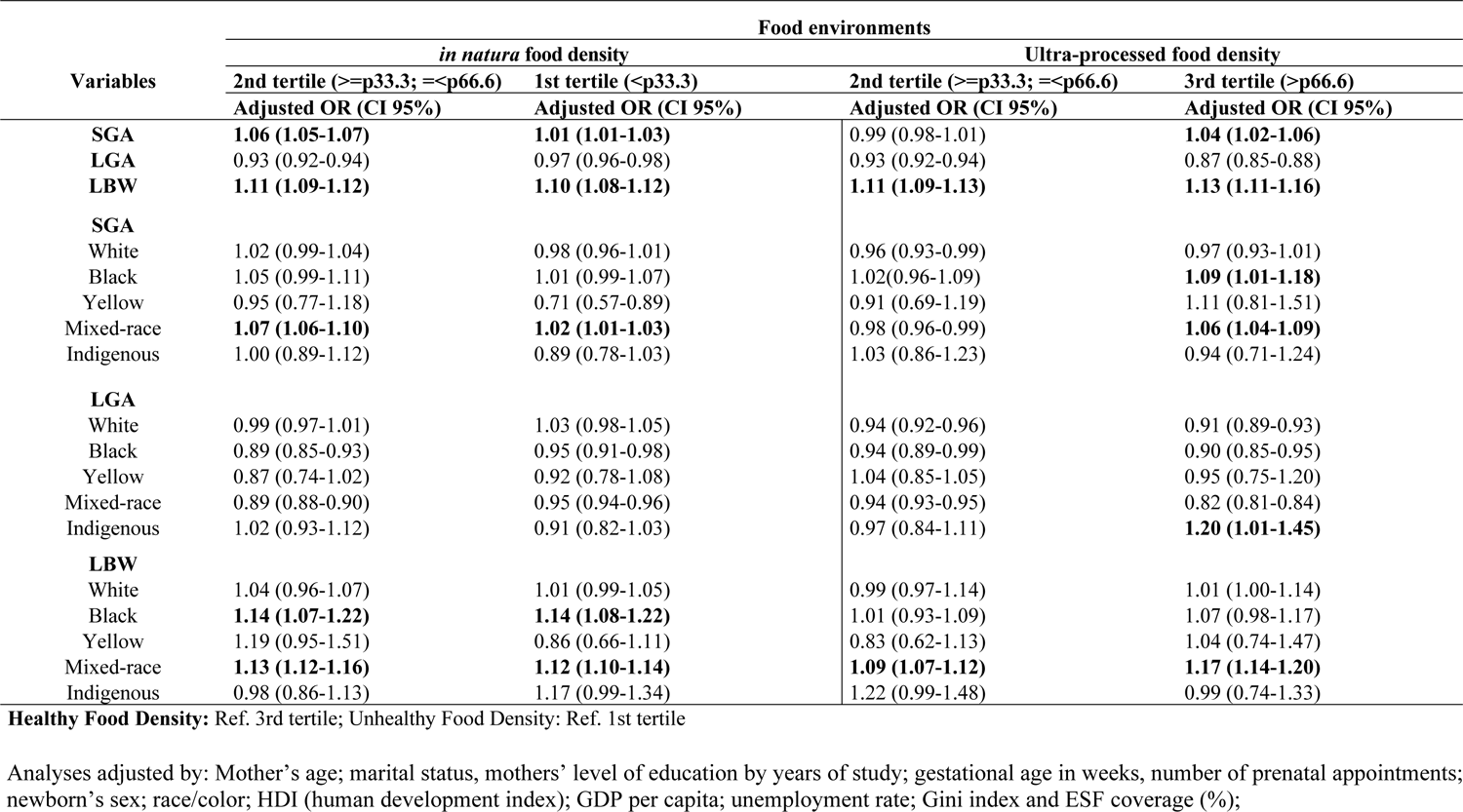
Adjusted Odds Ratio (OR) and 95% confidence interval for the association between food density and SGA, LGA, and LBW in binary logistic regression and models stratified by race (n=2,632,314).

## Discussion

Our objective was to study the association between food deserts, fetal growth, and birth weight. To the best of our knowledge, this is the first study to test this relation in Brazil. The results demonstrated that there are higher chances of women having an LBW and SGA newborn in municipalities with a higher supply of ultra-processed foods and low supply of *in natura* foods, mainly among black and mixed-race women. On the other hand, indigenous women who live in areas with a high density of ultra-processed foods have higher chances of having LGA newborns.

The results of this study support the position that the density of ultra-processed foods may play an even greater role than the density of *in natura* foods in fetal growth and birth weight; notably, the results were maintained after replicating the analyses, stratified by race. New data is that indigenous women have greater chances of LGA. Premature birth and SGA are considered the main causes of adverse neonatal outcomes worldwide.^22^

The association between food environments and fetal growth is a controversial issue. However, our study’s results align with the literature that considers that these environments may reinforce the effects of a poor diet, compromising perinatal and neonatal outcomes^13,22,24–27^. A similar study conducted by Sawangkum et al. (2020) revealed that women who lived in desert zones were associated with LBW newborns, and following s^24^. Limited access to food was associated with a SGA birth, following adjustment for maternal race/ethnicity^13^. Saeed and contributors in a study which aimed to evaluate the effect of maternal food insecurity on birth weight also demonstrated that women experiencing food insecurity had a greater risk of giving birth to a low weight newborn^26^. Maternal education, and having experienced a situation of food insecurity during pregnancy, were predictors of LBW^25^. Other studies also confirm this relation between limited access to food and the birth of LBW babies^28,29^. Research conducted with 8,753 households also demonstrated that the chance of having low weight babies was higher for those with food insecurity, in girls, and for mothers who had attended fewer prenatal appointments^30^. Equally, mothers in a situation of food insecurity had increased chances of giving birth to a LBW baby, mainly among females^31^. A different result concerning ultra-processed foods was observed in research where the percentage of energy intake of ultra-processed foods was a predictor of increased neonatal body fat^32^. This may explain the result observed in indigenous women living in municipalities with a higher density of ultra-processed foods and a higher chance of having LGA babies. This was previously associated with the invasion of the food industry in more rural zones, thereby changing the population’s consumption profile, and the lack of alternatives to access healthier food options. The industry is known to be the mediator for calorie increases, and consequent excessive gains in gestational weight^33–36^. Increased access to supermarkets is associated with less prevalence of overweight and obesity, and improved consumption of fruit and vegetables^27^.

Richterman, et al. 2020, observed that food insecurity was a risk factor for the birth of SGA babies^22^. The study analyzed 1,124,299 births, and noted that women with a low gain of gestational weight and who resided in food deserts, were at high risk of giving birth to premature babies, associated with other factors such as age and mother’s race, level of education, marital status and interpregnancy intervals^36^. The ecological study showed that women who lived in neighborhoods with food insecurity had higher chances of births of very SGA babies^37^.

Black women resident in regions with a lower supply of *in natura* foods have a greater risk of giving birth to low weight or premature newborns^38^. Risk markers and unfavorable birth outcomes related to limited access to healthy foods were more prevalent in areas of an average and high concentration of black people^39,40^. In our understanding, and as has been shown in various studies, we highlight the need for public social protection policies, considering social and health inequities related to the racial characteristics of minority and vulnerable social groups. The strong association with unfavorable outcomes for the conditions of being black, of mixed-race, and indigenous, can be observed in our results.

The main limitation of this study arises from the impossibility of attaining exposure (food density) in an individualized way to gain a better understanding of the context. However, the methodological design provided a practical approach to evaluate the objective of this study. Another possible limitation is the quality and reliability of secondary data, which may introduce biases related to missing values, under-estimation, and classification errors. In this respect, data was obtained from government sources, such as health information systems and the Ministry of Citizenship, which are known for holding high quality standards. There may also be some unobserved variables that confuse the association we have studied, such as the absence of variables on the mother’s nutritional status, and gains in gestational weight, or diet, for example, which are significant confounders for the topic studied.

## Conclusion

The study showed that there are greater chances of the birth of LBW and SGA babies among women from municipalities with a greater supply of ultra-processed foods, and low supply of *in natura* foods, mainly in black and mixed-race women. On the other hand, indigenous women who live in areas with a high density of ultra-processed foods have higher chances of giving birth to LGA babies, due to the greater supply of these unhealthy foods, which are of a low nutritional quality, and originate from unsustainable food systems. Thus, an urgent need is demonstrated for the creation of fairer and more egalitarian public policies in social terms, to guarantee access to healthy foods, which is a fundamental human right, to minimize these adverse perinatal and neonatal effects. Therefore, the timely provision of food supplements should be considered, through social assistance. This may help reduce perinatal and neonatal morbimortality, in addition to policies which range from promoting the production of agroecological and organic foods to using economic instruments and fiscal measures, such as taxation of food products with a high saturated fat, sugar, and salt content. Encouragement of food and nutritional educational activities, based on food guides for pregnant women should not be ruled out, to support and encourage the adoption of healthy habits. Future research should be encouraged, to understand this relation, and other maternal-infant outcomes.

## Data Availability

SINASC - https://pcdas.icict.fiocruz.br/conjunto-de-dados/sistema-de-informacao-sobre-nascidos-vivos/
CAISAN - https://aplicacoes.mds.gov.br/sagirmps/portal-san/artigo.php?link=23

## Acknowledgments

not applicable.

## Authorship

The authors’ contributions were as follows: A.V., M.D.V, N.J.S., and A.R.EG. designed the study; A.V., N.J.S., and A.F. collected data, and constructed the database; A.V., O.R.A, and R.C.R.S. outlined the analytical strategy; A.V., A.R.EG, M.M, S.P.X., and N.J.S. performed the statistical analyses, interpreted the results, and drafted the manuscript; A.R.EG, R.C.R.S., I.R.F., A.F., M.M, and P.R. interpreted the results and critically reviewed the manuscript. All of the authors have read the manuscript. We exclusively used public secondary and aggregated data. Therefore, informed consent and approval by the Research Ethics Committee was not required, in accordance with Resolution No. 466/2012 of the National Health Council of Brazil, National Research Ethics Commission.

## Data Availability

Data described in the manuscript, codebook, and analytical code, can be provided on request.

